# Uncovering Language Disparity of ChatGPT in Healthcare: Non-English Clinical Environment for Retinal Vascular Disease Classification

**DOI:** 10.1101/2023.06.28.23291931

**Authors:** Xiaocong Liu, Jiageng Wu, An Shao, Wenyue Shen, Panpan Ye, Yao Wang, Juan Ye, Kai Jin, Jie Yang

**Author notes:** ***Corresponding author* Prof. Jie Yang**, School of Public Health and The Second Affiliated Hospital, Zhejiang University School of Medicine, Address: 866 Yuhangtang Rd, Hangzhou, Zhejiang, China, 310058, **Prof. Kai Jin**, Eye Center, The Second Affiliated Hospital, School of Medicine, Zhejiang University, Hangzhou, Zhejiang, China. Address: 88 Jiefang Road, Hangzhou, China, 310009, **Prof. Juan Ye**, Eye Center, The Second Affiliated Hospital, School of Medicine, Zhejiang University, Hangzhou, Zhejiang, China. Address: 88 Jiefang Road, Hangzhou, China, 310009. Contribute equally to this paper (Xiaocong Liu, Jiageng Wu).

## Abstract

**Objective:** To evaluate the effectiveness and reasoning ability of ChatGPT in diagnosing retinal vascular diseases in the Chinese clinical environment.

**Materials and Methods:** We collected 1226 fundus fluorescein angiography reports and corresponding diagnosis written in Chinese, and tested ChatGPT with four prompting strategies (direct diagnosis or diagnosis with explanation and in Chinese or English).

**Results:** ChatGPT using English prompt for direct diagnosis achieved the best performance, with F1-score of 80.05%, which was inferior to ophthalmologists (89.35%) but close to ophthalmologist interns (82.69%). Although ChatGPT can derive reasoning process with a low error rate, mistakes such as misinformation (1.96%), and hallucination (0.59%) still exist.

**Discussion and Conclusions:** ChatGPT can serve as a helpful medical assistant to provide diagnosis under non-English clinical environments, but there are still performance gaps, language disparity, and errors compared to professionals, which demonstrates the potential limitations and the desiration to continually explore more robust LLMs in ophthalmology practice.

## INTRODUCTION

The global population of individuals with visual impairments exceeded 2.2 billion in 2019 and continues to rise^1^. As the leading causes of blindness, retinal vascular diseases are characterized by a complex array of clinical manifestations. Fundus fluorescein angiography (FFA) is a specialized ophthalmic test used to visualize the retinal vasculature^2^. However, interpreting the FFA results and making a diagnosis requires laborious analysis by experienced ophthalmologists. Recently, large language models (LLMs) like ChatGPT^3^, have demonstrated exceptional performance in various tasks due to their rich internal knowledge and strong deductive reasoning abilities^4–8^.

However, the related research within the medical field primarily focuses on the knwoledge assessment^9–12^, a comprehensive evaluation of ChatGPT’s capabilities in ophthalmology for disease diagnosis is lacking. Additionally, although existing LLMs demonstrate impressive cross-language understanding abilities, they may lead to significant disparities in non-English specific fields due to they were primarily trained on English corpus^13,14^. Therefore, in this study, by exploring ChatGPT’s ability to understand Chinese FFA reports, our objective is to evaluate ChatGPT’s diagnostic performance and inference abilities for retinal vascular diseases in a non-English clinical environment, and to find appropriate prompt strategies under these scenarios.

## MATERIALS AND METHODS

### Data Preparation

We collected 1226 Chinese FFA reports and the corresponding clinical diagnosis of 728 patients from the Eye Center of the Second Affiliated Hospital of Zhejiang University (SAHZU) between August 2016 and September 2021. The clinical diagnosis of each eye was either classified as *Normal* or one of the six primary retinal vascular diseases: diabetic retinopathy (DR), wet age-related macular degeneration (wetAMD), central serous chorioretinopathy (CSC), branch retinal vein occlusion (BRVO), central retinal vein occlusion (CRVO), and vogt-koyanagi-harada disease (VKH). The clinical diagnosis was based on a series of clinical information from the patients, primarily the FFA report. The patient data was de-identified, and all private information was removed. The approved IRB agreed to share access to the data with third parties, including sending it through APIs provided by companies like OpenAI, or using it in online platforms like ChatGPT.

### Diagnosis of Retinal Vascular Diseases Using ChatGPT

To diagnose the patient’s eye status based on FFA report with ChatGPT, we designed a fixed instruction that concatenates the patient’s FFA report as the whole prompt of ChatGPT. The instruction consists of a specific task description and all alternative conditoins. To fully exploit the potential of ChatGPT, we implemented different prompting strategies to investigate the potential effect and find the most appropriate way to apply it. First, we employed the *Direct* prompting strategy that requires ChatGPT to directly output the final option without explaination. Secondly, inspired by chain-of-thought prompting (CoT) technology^15^, we adopted *Step* prompting strategy to elicit the detailed reasoning process which provides interpretability for disease diagnosis. Finally, ChatGPT was primarily trained on the English corpus and may have difficulty in recognizing instructions and FFA reports in Chinese, as well as making use of internal knowledge. Therefore, we also rewrote the prompt in English while keeping the FFA reports in Chinese to conduct code-switching prompt examination. Therefore, we mainly investigate four prompt strategies: *Direct-cn, Step-cn, Direct-en and Step-en*. The detailed prompts can be found in the **Supplemental Table1**. Furthermore, we evaluated the robustness of prompting strategy by adopting different methods to conduct report analysis. All tests were conducted on the same version of GPT3.5-Turbo using the official API of OpenAI. **Figure 1** shows the overall workflow.

**Figure 1.**
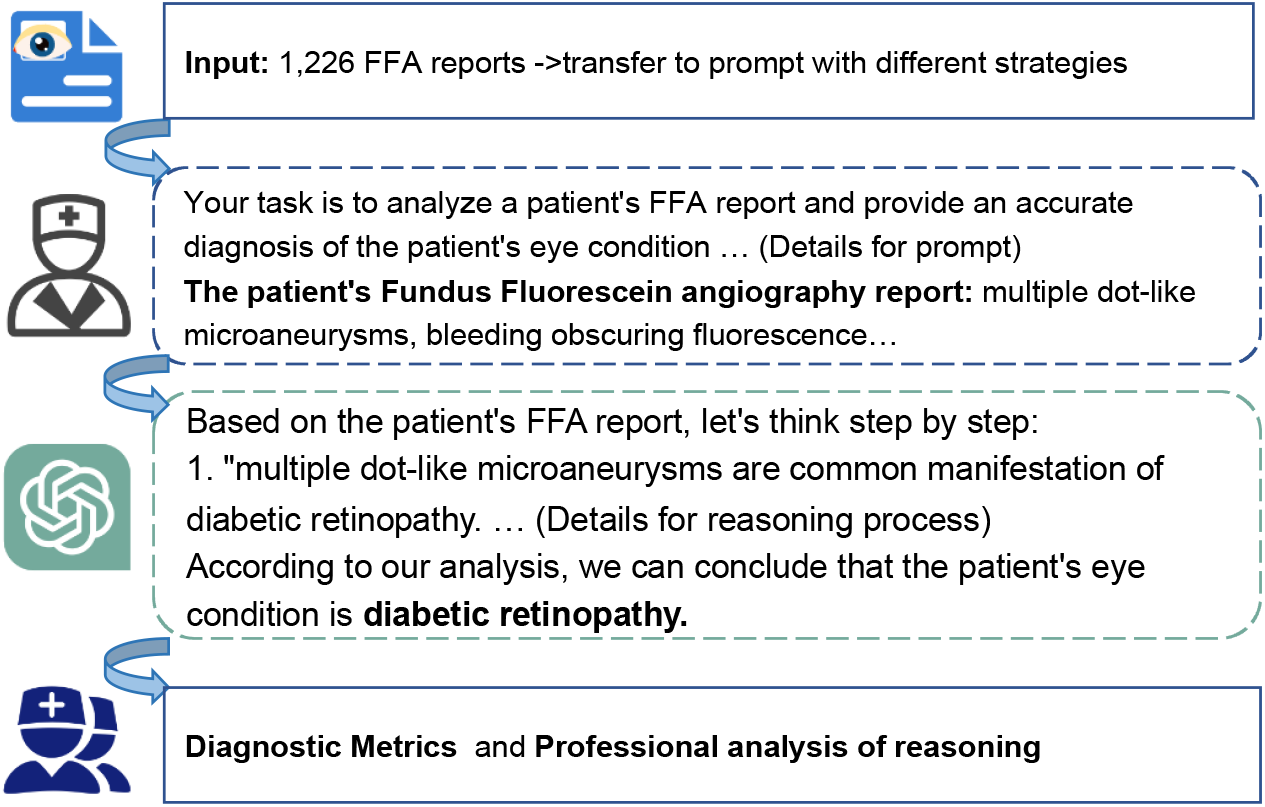
The Overall Workflow.

### Measurements and Definitions

We designed a systematic evaluation scheme to assess the performance of ChatGPT. In addition to diagnostic performance, we also incorporated a combination of inference ability, omission of information, hallucination, misinformation and inconsistency to evaluate the ChatGPT’s reasoning process.

1) Diagnostic performance: precision, recall and F1-score.

2) Inference ability: the total reasoning steps, the number of reasoning errors, and the incompleteness of the inference process.

3) Omission of information: whether crucial information from the original report was overlooked.

4) Hallucination: whether ChatGPT generates medical findings that were not present in the original report.

5) Misinformation: whether the output of ChatGPT quotes incorrect prior knowledge.

6) Inconsistency: whether the reasoning result is inconsistent with the reasoning process.

For diagnostic evaluation, precision, recall and F1-score were calculated based on the ChatGPT’s responses and gold clinical diagnosis. Additionally, to evaluate the diagnostic performance of ChatGPT, two ophthalmologists and two ophthalmology interns with two years of clinical experience from SAHZU were invited to diagnose 100 FFA reports, which were randomly selected according to the proportion of diseases.

In terms of the evaluation on ChatGPT’s inference ability, the last five measurements were evaluated on the responses of Step-cn and Step-en by two ophthalmologists from SAHZU. We randomly selected 509 FFA reports (no more than 100 for each disease) and the corresponding ChatGPT’s output for evaluation. Before the formal evaluation, ophthalmologists are requested to conduct annotation with training and the final inter-annotator agreement is up to 94%.

## RESULTS

The characteristics of FFA reports and responses of ChatGPT are listed in **Table 1**. Direct-cn and Direct-en directly gave the final options, and their mean (SD) lengths were 19.2 (4.4) and 5.7 (1.7), respectively, while Step-cn and Step-en provided detailed reasoning process, and their mean (SD) response lengths were 118.4 (71.8) and 100.5 (36.9), respectively. Examples of different prompts and their responses were recorded in the **Supplemental Table1**.

**Table 1.**
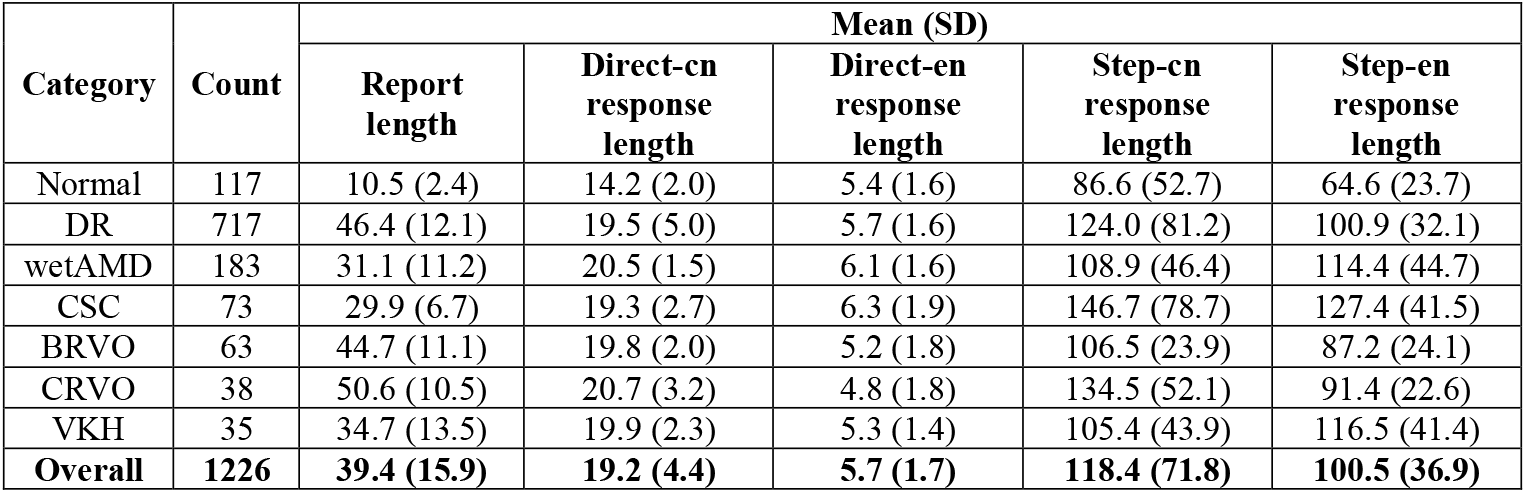
Characteristics of FFA Reports and ChatGPT Responses

| Category | Count | Mean (SD) |  |  |  |  |
| --- | --- | --- | --- | --- | --- | --- |
|  |  | Report length | Direct-cn response length | Direct-en response length | Step-cn response length | Step-en response length |
| Normal | 117 | 10.5 (2.4) | 14.2 (2.0) | 5.4 (1.6) | 86.6 (52.7) | 64.6 (23.7) |
| DR | 717 | 46.4 (12.1) | 19.5 (5.0) | 5.7 (1.6) | 124.0 (81.2) | 100.9 (32.1) |
| wetAMD | 183 | 31.1 (11.2) | 20.5 (1.5) | 6.1 (1.6) | 108.9 (46.4) | 114.4 (44.7) |
| CSC | 73 | 29.9 (6.7) | 19.3 (2.7) | 6.3 (1.9) | 146.7 (78.7) | 127.4 (41.5) |
| BRVO | 63 | 44.7 (11.1) | 19.8 (2.0) | 5.2 (1.8) | 106.5 (23.9) | 87.2 (24.1) |
| CRVO | 38 | 50.6 (10.5) | 20.7 (3.2) | 4.8 (1.8) | 134.5 (52.1) | 91.4 (22.6) |
| VKH | 35 | 34.7 (13.5) | 19.9 (2.3) | 5.3 (1.4) | 105.4 (43.9) | 116.5 (41.4) |
| **Overall** | **1226** | **39.4 (15.9)** | **19.2 (4.4)** | **5.7 (1.7)** | **118.4 (71.8)** | **100.5 (36.9)** |

### Diagnostic performance

The Direct-en achieved an overall precision of 79.61%, recall of 83.12%, and F1-score of 80.05%, which was 9.58% higher than that achieved by Direct-cn (**Table 2**). The diagnostic performance varied significantly for each disease. In these two prompts, ChatGPT performed better in the category of Normal and DR, with the F1-score exceeding 80%, but performed worse in the category of VKH and CSC, achieving F1-score of less than 4%. Additionally, the F1-score in the category of BRVO varied greatly, from 54.35% in Direct-cn to 74.51% in Direct-en.

**Table 2.** Performance of ChatGPT Across Various Disease Categories on FFA Reports

| Category | Direct-cn (%) |  |  | Direct-en (%) |  |  | Step-cn(%) |  |  | Step-en(%) |  |  |
| --- | --- | --- | --- | --- | --- | --- | --- | --- | --- | --- | --- | --- |
|  | P | R | F1 | P | R | F1 | P | R | F1 | P | R | F1 |
| Normal | 1.00 | 85.47 | 92.17 | 1.00 | 88.03 | 93.64 | 98.39 | 52.14 | 68.16 | 97.37 | 94.87 | 96.10 |
| DR | 91.55 | 72.52 | 80.93 | 91.05 | 95.12 | 93.04 | 85.07 | 95.40 | 89.94 | 82.13 | 93.58 | 87.48 |
| wetAMD | 44.72 | 87.98 | 59.30 | 59.92 | 80.87 | 68.84 | 63.58 | 60.11 | 61.80 | 60.00 | 34.42 | 43.75 |
| CSC | 4.35 | 2.74 | 3.36 | 33.33 | 1.37 | 2.63 | 34.15 | 19.18 | 24.56 | 50.00 | 6.85 | 12.05 |
| BRVO | 41.32 | 79.37 | 54.35 | 63.33 | 90.47 | 74.51 | 83.61 | 80.95 | 82.26 | 67.95 | 84.13 | 75.18 |
| CRVO | 93.10 | 71.05 | 80.60 | 84.85 | 73.68 | 78.87 | 41.27 | 68.42 | 51.49 | 58.33 | 73.68 | 65.12 |
| VKH | 0 | 0 | 0 | 0 | 0 | 0 | 0 | 0 | 0 | 0 | 0 | 0 |
| Overall | **75.03** | **70.15** | **70.47** | **79.61** | **83.12** | **80.05** | **76.24** | **77.16** | **75.61** | **74.56** | **75.94** | **73.46** |

In contrast, the Step-cn achieved an overall precision of 76.24%, recall of 77.16%, and F1-score of 75.61%, which was 2.15% higher than that achieved by ChatGPT with Step-en. Compared with Direct-cn, the F1-score of Step-cn was increased by 5.14%, and provided the reasoning process which is crucial for disease diagnosis. However, the diagnostic performance of Step-cn in the category of Normal and CRVO was far worse than that of Direct-cn. This is mainly because ChatGPT with Step-cn generates hallucinations for FFA reports of Normal category, which were wrongly diagnosed as CRVO. **Figure 2** further demonstrates the confusion matrix of ChatGPT with four prompting strategies.

**Figure 2.**
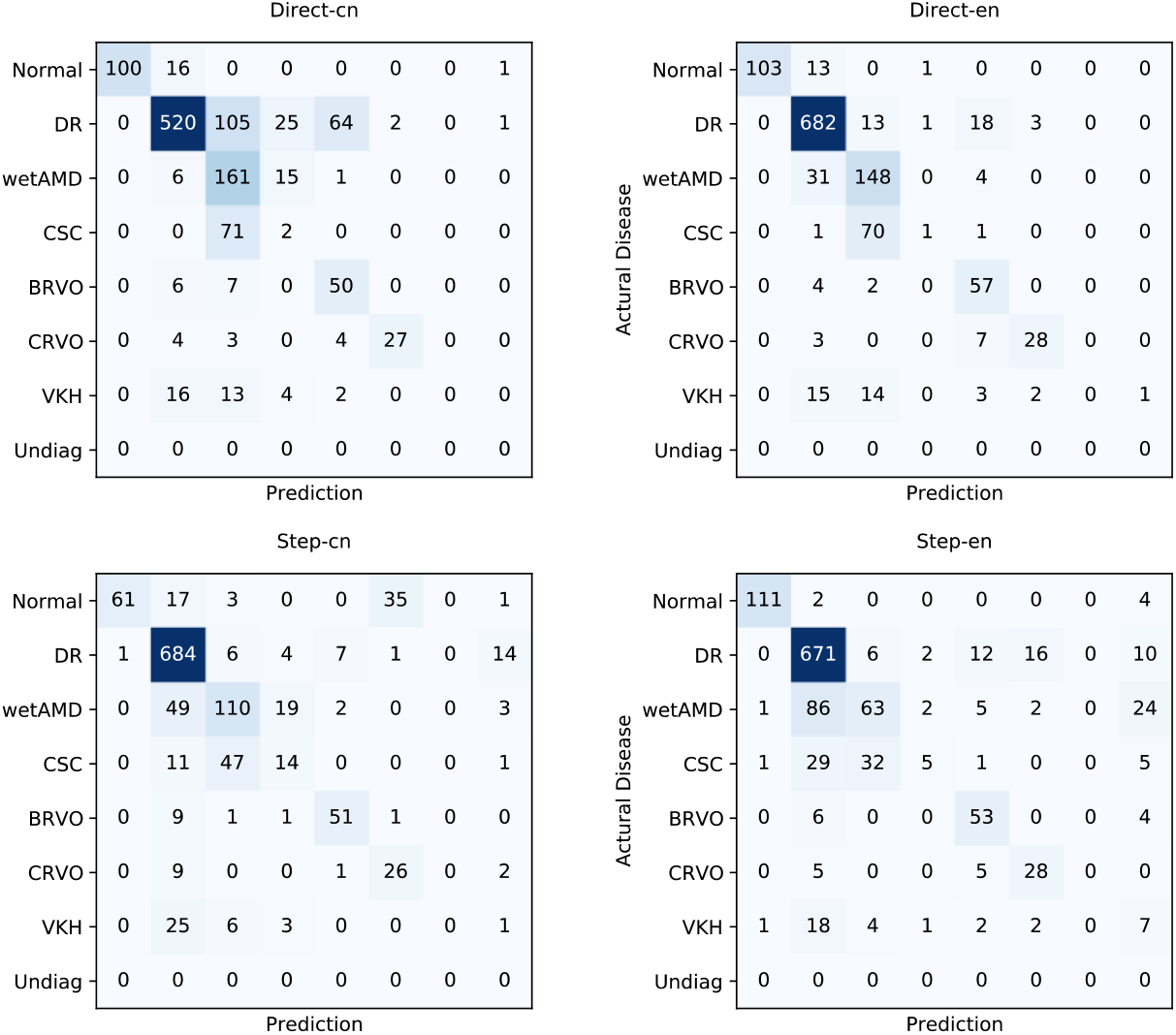
The Confusion Matrix of ChatGPT with Four Prompting Strategies.

**Figure 3(a)** shows the average F1-score of ophthalmologists, ophthalmology interns, ChatGPT with English prompts (Direct/Step-en) and Chinese prompts (Direct/Step-cn). Although ChatGPT performed better than experts in some disease types (e.g. Direct/Step-en in Normal and CRVO, Direct/Step-en and Direct/Step-cn in BRVO), the overall performance of ophthalmologists was the best (89.35%), followed by ophthalmology interns (82.69%), Direct/Step-en (76.76%) and Direct/Step-cn (73.04%).

**Figure 3.**
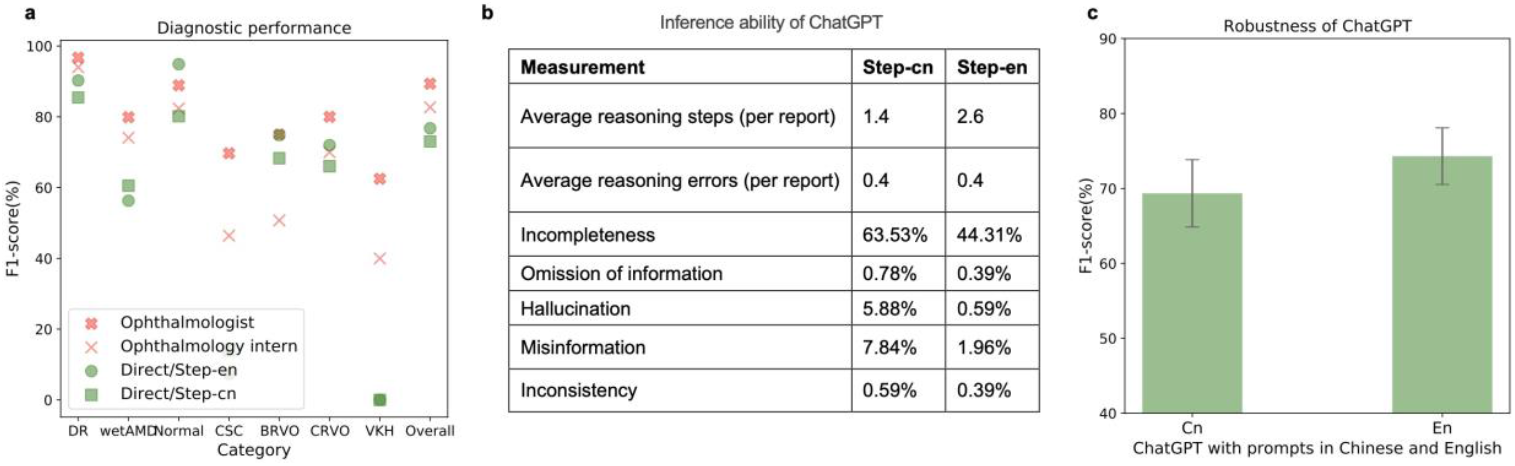
Diagnostic Performance of Human Beings and ChatGPT (a), Inference Ability of ChatGPT (b) and Robustness of ChatGPT (c)

### Inference ability

According to the assessment results of ophthalmologists in **Figure 3(b)**, in both Step-cn and Step-en, the average number of inference errors for each report was 0.4, but the latter brought less hallucination, misinformation and inconsistency. Instead, Step-cn, which involved fewer reasoning steps, was more prone to overlook the key information from the original report during the reasoning process. This led to an increase of 19.22% in the incompleteness of the inference process.

### Robustness

When using different prompting strategies, it is found that ChatGPT’s reply was not unique to any given FFA report. Hence, we evaluated the robustness of ChatGPT through various prompts in Chinese and English **(Supplemental Table1)**. As shown in **Figure 3(c)**, the mean (SD) F1-score of ChatGPT with Chinese and English prompt were 69.35% (4.49%) and 74.31% (3.78%), respectively. In a word, the diagnostic performance of ChatGPT with English prompts was better and more robust.

## DISCUSSION

To the best of our knowledge, this is the first study to evaluate ChatGPT’s performance on non-English clinical text for disease diagnosis. We have developed a systematic evaluation scheme that encompasses objective diagnostic performance, professional inference abilities, and comparisons with the diagnostic ability of experts.

Results demonstrated ChatGPT can preliminarily diagnose retinal vascular diseases based on Chinese FFA report and achieved a high F1-score of 80.05% at best. However, the diagnostic performance of ChatGPT varied significantly among different diseases and prompting strategies. The performance of common DR was significantly better than that of uncommon VKH, which is relatively low in incidence and more difficult to diagnose. The diagnostic performance of ChatGPT with English prompts was better and more robust than that of ChatGPT with Chinese prompts. Meanwhile, the diagnosis accompanied by reasoning steps was not necessarily lead to performance improvement, F1-scores decreased by 6.59% for English prompting and increased by 5.14% for Chinese prompting. This may be because ChatGPT was mainly trained on English corpus, and Direct-en facilitated a straightforward mapping from input to diagnosis, whereas Step-en trended to bring more mistakes than benefits through multi-step internal reasoning. But for Chinese prompts, the scarcity of Chinese training data results in limited knowledge for disease diagnosis. Step-cn with the requirement of a reasoning process can effectively compensate for incomplete and incorrect reasoning caused by limited knowledge, although it may introduce some noise. The performance gap between different diseases and prompting strategies demonstrates the potential unfairness brought by the overrepresentation of the major diseases, languages, and countries.

From the perspective of clinical practice, ChatGPT’s diagnostic performance still did not reach the level of ophthalmologists and even ophthalmology interns. It is worth noting that ChatGPT may be conservative in the diseases diagnosis. Despite the instruction restriction (must identy one), there were certain responses involved multiple conditions or indicated can not conclude based on existing information. Notably, although ChatGPT can derive reasoning process to improve clinical interpretability and interaction of clinical servive, ophthalmologists identified some harmful mistakes such as generating medical findings not mentioned in the original reports and quoting incorrect prior knowledge. More in-depth investigation and careful regulation are required before applying ChatGPT in the heathcare domain.

### Limitations

Our study has several limitations. Firstly, we did not fully ultilize the all information of clincal scenarios to conduct diagnosis, such as more detailed FFA images, which may reduce the diagnostic accuracy due to incomplete information. Since ChatGPT cannot analyse image, we will further evaluate the capabilities of multimodal models in subsequent research. Secondly, this study was not conducted in clinical practice. A prospective clinical trial can better examine LLM’s clinical benefit, we leave this to our future work.

## CONCLUSION

This study conducted extensive experiments to evaluate the diagnostic capabilities of ChatGPT in retinal vascular diseases, including objective diagnostic performance and professional reasoning analysis evaluated by ophthalmologists. ChatGPT with English prompts for direct diagnosis performed best, which was close to the diagnostic performance of ophthalmology interns with two years of clinical experience. On the contrary, due to limited Chinese training data and knowledge, ChatGPT with Chinese prompts led to incomplete reasoning and poor diagnostic performance, which demonstrates that there is a significant language disparity in the application of ChatGPT in clinical environment. Additionally, although ChatGPT can derive reasoning process with a low error rate, mistakes such as misinformation and hallucination still exist, which will mislead the diagnose of retinal vascular diseases. This study generally reveals the potential of LLMs to serve as an helpful medical assistant to provide diagnosis under non-English clinical environment, but also demonstrates the potential limitations and the desiration to continually explore more robust LLMs in ophthalmology practice.

## AUTHOR CONTRIBUTIONS

XL and JW conducted experiments, statistical analysis, and drafting the work, and contributed equally to this work as co–first authors. Jie Y designed this study. AS and WS participated in the data extraction. AS, WS, PY and YW participated in the expert evaluation. Juan Y, KJ and Jie Y are corresponding authors, providing administrative, technical, and material support. All authors revised the manuscript and approved the submitted version.

## DATA AVAILABILITY

Data will be made available for research purposes upon request. Data requests are to be directed to.

## REFERENCES

1. Bourne R, Steinmetz JD, Flaxman S, et al. Trends in prevalence of blindness and distance and near vision impairment over 30 years: an analysis for the Global Burden of Disease Study. The Lancet Global Health 2021;9(2):e130–e143

2. Marmoy OR, Henderson RH, Ooi K. Recommended protocol for performing oral fundus fluorescein angiography (FFA) in children. Eye 2022;36(1):234–236.

3. ChatGPT: Optimizing Language Models for Dialogue. https://openai.com/blog/chatgpt/. Accessed Accessed June 26, 2023.

4. King MR. The future of AI in medicine: a perspective from a chatbot. Ann Biomed Eng 2023;51(2):291–295.

5. Liu Y, Han T, Ma S, et al. Summary of ChatGPT/GPT-4 research and perspective towards the future of large language models. Published online First: 4 April 2023. https://arxiv.org/abs/2304.01852. Accessed June 26, 2023.

6. Janssen BV, Kazemier G, Besselink MG. The use of ChatGPT and other large language models in surgical science. BJS Open 2023;7(2):zrad032.

7. Liu S, Wright AP, Patterson BL, et al. Using AI-generated suggestions from ChatGPT to optimize clinical decision support. JAMIA 2023;30(7):1237–45.

8. Jiang LY, Liu XC, Nejatian NP, et al. Health system-scale language models are all-purpose prediction engines. Nature 2023. doi: 10.1038/s41586-023-06160-y.

9. Kung TH, Cheatham M, Medenilla A, et al. Performance of ChatGPT on USMLE: potential for AI-assisted medical education using large language models. PLOS Digit Health 2023;2(2):e0000198.

10. Kumah-Crystal Y, Mankowitz S, Embi P, et al. ChatGPT and the clinical informatics board examination: the end of unproctored maintenance of certification? JAMIA 2023;ocad104.

11. Mihalache A, Popovic MM, Muni RH. Performance of an artificial intelligence chatbot in ophthalmic knowledge assessment. JAMA Ophthalmol 2023; 141(6):589–597.

12. Sarraju A, Bruemmer D, Van Iterson E, et al. Appropriateness of cardiovascular disease prevention pecommendations obtained from a popular online chat-based artificial intelligence model. JAMA 2023;329(10):842.

13. Lai VD, Ngo NT, Veyseh APB, et al. ChatGPT beyond English: towards a comprehensive evaluation of large language models in multilingual learning. Published online First: 12 April 2023. https://arxiv.org/abs/2304.05613. Accessed June 26, 2023.

14. Ferrara E. Should ChatGPT be biased? challenges and risks of bias in large language models. Published online First: 7 April 2023. https://arxiv.org/abs/2304.03738. Accessed June 26, 2023.

15. Wei J, Wang X, Schuurmans D, et al. Chain-of-Thought prompting elicits reasoning in large language models. Published online First: 28 January 2022. https://arxiv.org/abs/2201.11903. Accessed June 26, 2023.

