## Supplemental Table1 for "Uncovering Language Disparity of ChatGPT in Healthcare: Non-English Clinical Environment for Retinal Vascular Disease Classification"

**Supplemental Table1. Example of Input and Output of ChatGPT with Various Prompts**

| **Prompt strategy** | **Input** | **Output** | **Gold Standard** |
| --- | --- | --- | --- |
| Direct-cn | ***Translation：****Your task is to determine which of the following eye conditions the patient has: (A) Normal, (B) Diabetic retinopathy, (C) Age-related macular degeneration, (D) Central serous chorioretinopathy, (E) Branch retinal vein occlusion, (F) Central retinal vein occlusion, (G) Vogt Koyanagi Harada Syndrome. Please output the only correct option of eye condition.*  *The patient's Fundus Fluorescein angiography report: In the left eye, mottled fluorescence is observed in the macular area. There is a small area of bleeding in the center that blocks the fluorescence. Peripheral areas show dot-like high fluorescence. In the later stages, there is leakage of fluorescence in the macular area.* | ***Translation：****The patient has (C) Age-related macular degeneration.* | Age-related macular degeneration |
| Direct-en | Your task is to determine which of the following eye conditions the patient has: (A) Normal, (B) Diabetic retinopathy, (C) Age-related macular degeneration, (D) Central serous chorioretinopathy, (E) Branch retinal vein occlusion, (F) Central retinal vein occlusion, (G) Vogt Koyanagi Harada Syndrome. Please output the only correct option of eye condition.  The patient's Fundus Fluorescein angiography report: *(****Translation：****In the left eye, mottled fluorescence is observed in the macular area. There is a small area of bleeding in the center that blocks the fluorescence. Peripheral areas show dot-like high fluorescence. In the later stages, there is leakage of fluorescence in the macular area.* | The patient has (C) Age-related macular degeneration. | Age-related macular degeneration |
| Step-cn | ***Translation：****Your task is to analyze a patient's Fundus Fluorescein angiography report and determine exactly which of the following eye condition the patient is: (A) Normal, (B) Diabetic retinopathy, (C) Age-related macular degeneration, (D) Central serous chorioretinopathy, (E) Branch retinal vein occlusion, (F) Central retinal vein occlusion, (G) Vogt Koyanagi Harada Syndrome. The final diagnosis must be limited to one option from the given eight options and cannot involve multiple selections at once. Output should be structured as following format: "Based on the patient's Fundus Fluorescein angiography report, Let's think step by step: {reasoning process}. According to our comprehensive analysis, we can conclude that the patient's eye condition is {eye condition option}."*  *The patient's Fundus Fluorescein angiography report: The left eye shows scattered microaneurysms in the retina, capillary dilation and leakage, small non-perfused areas, late-phase fluorescence leakage, and cystoid accumulation in the macular region.)* | ***Translation：****We analyzed the patient's fundus fluorescein angiography report step by step. The report indicates the presence of scattered microaneurysms, capillary dilation and leakage, small non-perfused areas, late-phase fluorescein leakage, and cystoid macular edema in the left eye. These symptoms suggest that the patient may have diabetic retinopathy since microaneurysms and capillary dilation and leakage are typical manifestations of diabetic retinopathy. Non-perfused areas and fluorescein leakage are also common symptoms of this condition. The presence of cystoid macular edema suggests that the patient may have macular involvement, which is a common complication of diabetic retinopathy. Therefore, we can conclude that the patient's ocular condition is diabetic retinopathy.* | Diabetic retinopathy |
| Step-en | Your task is to analyze a patient's Fundus Fluorescein angiography report and determine exactly which of the following eye condition the patient is: (A) Normal, (B) Diabetic retinopathy, (C) Age-related macular degeneration, (D) Central serous chorioretinopathy, (E) Branch retinal vein occlusion, (F) Central retinal vein occlusion, (G) Vogt Koyanagi Harada Syndrome. The final diagnosis must be limited to one option from the given eight options and cannot involve multiple selections at once. Output should be structured as following format: "Based on the patient's Fundus Fluorescein angiography report, Let's think step by step: {reasoning process}. According to our comprehensive analysis, we can conclude that the patient's eye condition is {eye condition option}."  The patient's Fundus Fluorescein angiography report: *(****Translation****：The left eye shows scattered microaneurysms in the retina, capillary dilation and leakage, small non-perfused areas, late-phase fluorescence leakage, and cystoid accumulation in the macular region.)* | Based on the patient's Fundus Fluorescein angiography report, we can see that there are scattered microaneurysms in the left retina, as well as capillary dilation and leakage. There are also small areas of non-perfusion and late-stage fluorescence leakage, as well as cystic accumulation in the macular area. These findings are indicative of Diabetic retinopathy. Therefore, according to our comprehensive analysis, we can conclude that the patient's eye condition is Diabetic retinopathy. | Diabetic retinopathy |
| Step-cn (more detail) | ***Translation****：As an experienced clinical ophthalmologist. Your task is to analyze a Chinese patient's Fundus Fluorescein angiography (FFA) report and determine his/her eye condition. The patient may have a normal eye (A), or one of the following eye diseases: (B) Diabetic retinopathy, (C) Age-related macular degeneration, (D) Central serous chorioretinopathy, (E) Branch retinal vein occlusion, (F) Central retinal vein occlusion, (G) Vogt Koyanagi Harada Syndrome.*  *To accomplish this task, you will need to conduct an in-depth analysis of each manifestation in the report, using medical knowledge and scientific facts to guide your reasoning process. As you progress through your analysis, be sure to explain each step of your reasoning process in detail, allowing patients to easily understand how you arrived at your conclusion. Although sometimes it may be difficult to reach an exact diagnosis from this report alone, you should also select the most likely eye condition rather than saying cannot diagnose. Finally, be sure to find as much reliable evidence as possible to support your reasoning process. Your answers should be flexible enough to provide various relevant and creative explanations for reasoning process.*  *In your response, please follow the structure below: 'Based on the patient's Fundus Fluorescein angiography report, Let's think step by step: {reasoning process}.*  *According to our comprehensive analysis, we can conclude that the patient's eye condition is {eye condition option}.'*  *The patient's Fundus Fluorescein angiography report:* *Left eye: Hemorrhage adjacent to the optic disc with cotton wool spot formation, followed by vascular leakage.* | ***Translation：****According to the patient's Fundus Fluorescein angiography report, let's consider the findings step by step:*  *Hemorrhage adjacent to the optic disc in the left eye: This means that there is bleeding next to the optic disc in the left eye, which may be due to vascular rupture or inflammation. This condition may be associated with various eye diseases.*  *Formation of cotton wool spots: Cotton wool spots are white or gray patches typically associated with retinal artery or vein occlusion. This condition may be related to branch retinal vein occlusion or central retinal vein occlusion.*  *Late-phase vascular leakage: This means that in the late phase, fluorescein dye leaks from the blood vessels into the surrounding tissues. This condition may be associated with diabetic retinopathy, age-related macular degeneration, central serous chorioretinopathy, or Vogt-Koyanagi-Harada syndrome.*  *Based on our comprehensive analysis, we can conclude that the patient's ocular condition is consistent with branch retinal vein occlusion. This is supported by the presence of hemorrhage adjacent to the optic disc and the formation of cotton wool spots, which are typical manifestations of branch retinal vein occlusion. The late-phase vascular leakage also supports this diagnosis. Although other eye diseases may also cause similar symptoms, branch retinal vein occlusion is the most likely diagnosis.* | Branch retinal vein occlusion |
| Step-en (more detail) | As an experienced clinical ophthalmologist. Your task is to analyze a Chinese patient's Fundus Fluorescein angiography (FFA) report and determine his/her eye condition. The patient may have a normal eye (A), or one of the following eye diseases: (B) Diabetic retinopathy, (C) Age-related macular degeneration, (D) Central serous chorioretinopathy, (E) Branch retinal vein occlusion, (F) Central retinal vein occlusion, (G) Vogt Koyanagi Harada Syndrome.  To accomplish this task, you will need to conduct an in-depth analysis of each manifestation in the report, using medical knowledge and scientific facts to guide your reasoning process. As you progress through your analysis, be sure to explain each step of your reasoning process in detail, allowing patients to easily understand how you arrived at your conclusion. Although sometimes it may be difficult to reach an exact diagnosis from this report alone, you should also select the most likely eye condition rather than saying cannot diagnose. Finally, be sure to find as much reliable evidence as possible to support your reasoning process. Your answers should be flexible enough to provide various relevant and creative explanations for reasoning process.  In your response, please follow the structure below: 'Based on the patient's Fundus Fluorescein angiography report, Let's think step by step: {reasoning process}.  According to our comprehensive analysis, we can conclude that the patient's eye condition is {eye condition option}.'  The patient's Fundus Fluorescein angiography report: *(****Translation****：Left eye: Hemorrhage adjacent to the optic disc with cotton wool spot formation, followed by vascular leakage.)* | Based on the patient's Fundus Fluorescein angiography report, let's think step by step:  1. the report is referring to the patient's left eye.  2. there is bleeding near the optic disc, which is the area where the optic nerve enters the eye.  3. the presence of cotton wool spots, which are white patches on the retina caused by damage to nerve fibers.  4. there is leakage of blood vessels in the later stages of the angiography.  According to our comprehensive analysis, we can conclude that the patient's eye condition is most likely Diabetic Retinopathy (B). Diabetic Retinopathy is a complication of diabetes that affects the blood vessels in the retina, leading to bleeding, cotton wool spots, and leakage of blood vessels. The presence of bleeding near the optic disc and cotton wool spots in the patient's left eye, along with the later-stage leakage of blood vessels, are all consistent with the diagnosis of Diabetic Retinopathy. However, further examination and testing may be necessary to confirm the diagnosis. | Branch retinal vein occlusion |
| Step-cn (two steps) | ***Translation****：According to a patient's Fundus Fluorescein angiography report and its analysis, your task is to determine exactly which of the following eye condition the patient is: (A) Normal, (B) Diabetic retinopathy, (C) Age-related macular degeneration, (D) Central serous chorioretinopathy, (E) Branch retinal vein occlusion, (F) Central retinal vein occlusion, (G) Vogt Koyanagi Harada Syndrome. The final diagnosis must be limited to one option from the given seven options and cannot involve multiple selections at once. Please output the only correct option of eye condition. The patient's Fundus Fluorescein angiography report: Right eye: Central retinal vein occlusion, delayed filling of retinal veins, scattered patchy hemorrhages causing fluorescence blockage, small avascular areas, and capillary dilation with leakage.*  *Analysis: Let's think step by step. The report states that the patient has "central retinal vein occlusion in the right eye." This rules out options A, B, C, D, E, and G, leaving us with option F, central retinal vein occlusion, as the final diagnosis.*  *Diagnosis: "* | ***Translation****：Central retinal vein occlusion* | Central retinal vein occlusion |
| Step-en (two steps) | According to a patient's Fundus Fluorescein angiography report and its analysis, your task is to determine exactly which of the following eye condition the patient is: (A) Normal, (B) Diabetic retinopathy, (C) Age-related macular degeneration, (D) Central serous chorioretinopathy, (E) Branch retinal vein occlusion, (F) Central retinal vein occlusion, (G) Vogt Koyanagi Harada Syndrome. The final diagnosis must be limited to one option from the given seven options and cannot involve multiple selections at once. Please output the only correct option of eye condition. The patient's Fundus Fluorescein angiography report: *(****Translation****：Right eye: Central retinal vein occlusion, delayed filling of retinal veins, scattered patchy hemorrhages causing fluorescence blockage, small avascular areas, and capillary dilation with leakage.)*  Analysis: Let's think step by step. The report states that the patient has "central retinal vein occlusion in the right eye." This rules out options A, B, C, D, E, and G, leaving us with option F, central retinal vein occlusion, as the final diagnosis.  Diagnosis: " | Central retinal vein occlusion | Central retinal vein occlusion |

* According to medRxiv policies, Chinese text has been replaced by the English translation. The original text can be made available upon request.
